# Maternal diagnoses around pregnancy and risk of autism spectrum disorder – A population-based study

**DOI:** 10.1101/2022.06.28.22276974

**Authors:** Vahe Khachadourian, Elias Arildskov, Jakob Grove, Paul F O’Reilly, Arad Kodesh, Stephen Z Levine, Joseph D Buxbaum, Abraham Reichenberg, Lisa A. Croen, Diana Schendel, Sven Sandin, Stefan Nygaard Hansen, Magdalena Janecka

## Abstract

**Background:** There is growing interest in the potential effects of maternal health in pregnancy on the risk for autism spectrum disorder (ASD) in the offspring. Studies to date have examined only a few maternal diagnoses in pregnancy and did not account for the potential effects of co-occurring conditions. This study systematically investigates the associations between maternal diagnoses in pregnancy and the risk of ASD in the offspring, addressing comorbidity and familial confounding.

**Methods:** The study sample included all children born in Denmark in 1998-2007 (N=653,580) and their mothers. We fitted Cox proportional hazard regression models to estimate the risk of ASD associated with maternal chronic and non-chronic ICD-10 diagnoses. We further examined the evidence for potential familial confounding using discordant sibling and negative control designs.

**Results:** The analytic sample included 7,866 ASD cases (1.2%) and 645,714 controls, born to 425,399 mothers. We tested 241 non-chronic, and 90 chronic maternal conditions for an association with ASD. In a multivariable model, adjusting for maternal comorbidities, 15 of the non-chronic and 5 of the chronic diagnoses were significantly associated with ASD after correcting for multiple testing. Pregnancy complications and injuries accounted for a major portion of the non-chronic maternal diagnoses associated with risk of ASD (e.g., diabetes mellitus in pregnancy: HR=1.27, 95%CI: 1.07-1.51; fracture of skull and facial bones: HR=1.61, 95%CI: 1.09-2.38), and maternal psychiatric diagnoses were most strongly associated with ASD among the chronic conditions (e.g., major depressive disorder: HR=1.89, 95%CI: 1.39, 2.56). Results from negative control (timing of diagnosis, before vs. during pregnancy) and sibling analyses provide limited evidence for the role of familial confounding in the observed associations.

**Conclusions:** Our findings highlight the potential relevance of maternal health around pregnancy for offspring long-term neurodevelopmental outcomes and offer insights into the ASD etiology – necessitating further dissection of the mechanisms underlying the observed associations.

## INTRODUCTION

Autism spectrum disorder (ASD) is a neurodevelopmental disorder characterized by impairments in social communication and restrictive and repetitive behaviors. Due to the early neurodevelopmental origins of ASD, studies on non-genetic risk factors for the disorder have focused on maternal exposures during pregnancy^1^.

To date, epidemiological studies have documented associations between offspring ASD and several maternal medical conditions, occurring around pregnancy including depression^2^, diabetes^3^, immune system diseases^4^, and infections^5^. However, pregnant women experience many other health problems, most of which have not been studied in the relation of ASD risk in the offspring. Furthermore, in the light of frequent co-occurrence of medical conditions in pregnancy^6^, the degree to which the association of a specific maternal condition with ASD may be due to their co-occurrence with other diagnoses remains unknown. Finally, mechanistic interpretations of the observed associations remain challenging. Maternal medical diagnoses recorded in health registers may be associated with adverse effects stemming from having the disorder (e.g., possible changes in maternal physiology, medication use) or associated with factors linked to the family, including family residence in polluted areas, parental socioeconomic status, or an underlying genetic predisposition shared by family members.

Various study designs can be implemented to distinguish between familial and non-familial factors in observational data. For instance, sibling designs can control for family level and sibling-invariant confounders without the need to specify, measure, or define the functional form of these variables^7^. The use of negative controls – e.g., paternal exposures during pregnancy or maternal exposures during a non-pregnancy period - is another approach that can be applied to identify unmeasured familial confounding linked to maternal exposures and other types of biases^8^. Thus, complementing standard observational study designs with the use of these specialized study designs will be critical to providing insights into mechanisms underlying ASD etiology, and the degree to which the identified risk factors are modifiable.

Here we comprehensively assess maternal diagnoses and their associations with offspring ASD, accounting for comorbidity and chronicity of the conditions using a large, population-based cohort study from Denmark. Furthermore, we complement our cohort study approach with other study designs in order to assess the impact of unmeasured familial confounding on the observed associations.

## METHODS

### Study design and population

The source population for the study included all children born between January 1^st^, 1998 and December 31^st^, 2007 in Denmark and their mothers identified in the Danish Central Population Register (DCPR)^9–11^

### Outcome

Diagnosis of ASD (ICD-10: F84.0, F84.1, F84.5, F84.8, F84.9, and their subcodes), was obtained via linkage with the Danish Psychiatric Central Research Register (DPCRR)^10^ and Danish National Patient Register (DNPR)^9^. In Denmark, general practitioners or school psychologists refer children with a suspected ASD to a child and adolescent psychiatric department where they undergo a multidisciplinary evaluation. All ASD conditions are diagnosed by child and adolescent psychiatrists and are reported according to the International Classification of Disease-version 10 (ICD-10) since 1994; diagnoses from both in- and outpatient contacts have been reported to the DPCRR since 1995. All children in the sample were followed from birth until the first diagnosis of ASD, death, or end of follow-up (December 31, 2012), whichever occurred first^12^.

### Exposure

Maternal diagnoses were obtained via linkage with the Psychiatric Central Research Register (diagnoses of mental disorders) and National Patient Register (diagnoses of non-mental conditions). All reported maternal diagnoses occurring during the exposure detection window (see below) and coded using the ICD-10 (i.e., any reported diagnoses since 1994) served as the exposures. The hierarchical organization of diagnostic codes in ICD-10 has 4 levels, presenting information from least to most specific diagnosis. The current analyses were based on level 3 diagnostic codes (hereafter referred to as “diagnoses” for brevity), e.g., F33 for major depressive disorder. We used the Chronic Condition Indicator developed by the Agency for Healthcare Research and Quality^13^ to assign the diagnoses into chronic and non-chronic categories. Any level 3 diagnostic category that included both chronic and non-chronic sub-diagnoses at level 4 were assessed and classified on a case-by-case basis by clinical experts in the team. All exposure variables were binary indicators of presence / absence of the given diagnosis in the exposure period. All diagnoses present in the register but not corresponding to any of the ICD-10 codes were omitted.

#### Detection window

##### Non-chronic diagnoses

in the main analyses the detection period for non-chronic diagnoses comprised the 24 months preceding the childbirth. Assuming a full-term pregnancy, the 24-month period captured the pregnancy and the 15 months preceding conception. Using a relatively wide detection period was dictated by the intention to capture non-chronic diagnoses occurring during pregnancy as well as those occurring before pregnancy, but whose residual effects may still potentially affect fetal development.

##### Chronic diagnoses

in the main analyses the detection period for chronic diagnoses was defined as the 48 months preceding the childbirth. Widening of the exposure window compared to non-chronic diagnoses was dictated by the assumption that chronic diagnoses are permanent following their onset, and the onset timing cannot be precisely determined using the registry-based data. In this situation, the wider detection window for chronic diagnoses provided a higher sensitivity in capturing maternal chronic diagnoses that may not be entered into the registry around pregnancy (e.g., well-managed conditions that do not require frequent medical attention).

### Covariates

The study covariates included child’s sex and year of birth, maternal age at childbirth, and the total number of new and repeated maternal diagnoses, both chronic and non-chronic, during the 24 months preceding childbirth. Birth year was intended to account for varying incidence of ASD over time^14^; maternal age accounted for maternal age differences in ASD risk^15,16^ and in risk for medical condition during pregnancy^17^; and accounting for the total number of maternal diagnoses during the 24 months preceding childbirth was considered as a proxy measure for health-seeking behaviors and healthcare utilization and was included in all models. Covariate information was obtained from Danish Central Population Register (DCPR), DPCRR^10^, and DNPR^9^

### Statistical Analysis

The analyses comprised two phases. The first phase systematically tested the associations between each maternal diagnosis and ASD and the second evaluated the impact of unmeasured familial factors on these associations with non-chronic conditions. In the second phase we employed two orthogonal approaches (i) contrasting the effects on ASD risk of maternal diagnoses around pregnancy (within the 24-month detection window) to the risk effects of those diagnoses reported more temporarily distal to pregnancy, and (ii) contrasting ASD risk in siblings exposed and non-exposed to the same maternal diagnosis. These approaches may not provide reliable inference for the *chronic* diagnoses due to the limitations in ascertaining the disease onset, and potential misclassification of exposed/unexposed siblings.

#### Phase 1: Associations between maternal diagnoses and ASD

In phase 1 analyses we evaluated associations between maternal diagnoses and risk of ASD in offspring. First, each maternal diagnosis was tested separately for an association with ASD using a Cox proportional hazard model implemented in the *survival* package^18^, adjusting for the study covariates. Clustering sandwich estimators were used to account for within-family correlations occurring due to the presence of siblings in the dataset. All maternal diagnoses which occurred in <20 mothers of children with ASD and/or mothers of children without ASD were excluded from the analyses to minimize the risk of sparse data bias^19^. The statistical significance of these individual associations was assessed using P-values adjusted for false discovery rates (q-value)^20^ at the q-value threshold of 0.05.

Next, to adjust for possible correlation (i.e., co-occurrence) between different diagnoses, all diagnoses with a statistically significant association with ASD in individual adjusted models (q-value < 0.05), including both chronic and non-chronic diagnoses, were concurrently included in a multivariable model, adjusting for all covariates.

#### Phase 2: Assess impact of unmeasured familial confounding on maternal non-chronic diagnoses and ASD associations

All Phase 2 analyses to assess impact of unmeasured familial confounding were performed only for the non-chronic diagnoses with a statistically significant association with offspring ASD in the Phase 1 analyses after controlling for multiple testing using false discovery rates adjustment.

#### Timing of maternal diagnosis

We repeated the multivariable analyses described in the Phase 1 analyses section, additionally including non-chronic diagnostic categories recorded more distally to the index childbirth (24 to 48 months before the index childbirth, including diagnostic codes related to previous pregnancies). This allowed us to compare associations between non-chronic maternal diagnoses and offspring ASD by their timing, accounting for the co-occurring conditions and covariates.

#### Discordant sibling analysis

To assess the effect of time-invariant familial confounders on the observed associations, we repeated the primary multivariable model, treating maternal IDs as a stratification variable in the Cox regression model, restricting the sample to children with at least one sibling. We repeated the multivariable model from the primary analysis in this subsample to allow for direct comparison of the results between sibling and Phase 1 (multivariable) analyses.

All analyses were implemented in R software (version 4.0.3)^21^. Evidence for familial confounding in the association between specific maternal non-chronic diagnoses around pregnancy and ASD was inferred from either (i) non-null associations of non-chronic diagnoses 24-48 months before the childbirth (negative exposure controls) with ASD; or (ii) differences between the non-chronic condition risk estimates from the Phase 1 and discordant sibling analyses.

## RESULTS

The study sample included 653,580 children born in Denmark to 425,399 mothers between 1998 and 2007. A total of 7,866 children received an ASD diagnosis (1.2%) during the study follow-up period. The median duration of follow-up was 9.9 (IQR: 7.4 – 12.5) years. The median age at first ASD diagnosis was 6.8 years (IQR: 4.8 – 9.4). Maternal age at childbirth did not differ by child ASD status (with/without ASD: 30.1 years vs. 30.3 years) nor did the mean number of maternal diagnoses during the 2 years before the childbirth (9.9 vs. 9.3). Table 1 presents the sample characteristics by ASD diagnosis.

**Table 1.** Demographic characteristics of the analytical sample (mother-child dyads)

| **Variables** | **ASD**<br>**( n = 7866)** | **No ASD**<br>**( n = 645,714)** | **Total**<br>**(N = 653,580)** |
| --- | --- | --- | --- |
| Maternal age at delivery, mean (SD) | 30.1 (5.1) | 30.3 (4.8) | 30.3 (4.8) |
| Child's year of birth, n (%) |  |  |  |
| 1998 | 1098 (14.0%) | 65162 (10.1%) | 66260 (10.1%) |
| 1999 | 1126 (14.3%) | 65186 (10.1%) | 66312 (10.1%) |
| 2000 | 1105 (14.0%) | 66071 (10.2%) | 67176 (10.3%) |
| 2001 | 935 (11.9%) | 64511 (10.0%) | 65446 (10.0%) |
| 2002 | 856 (10.9%) | 63252 (9.8%) | 64108 (9.8%) |
| 2003 | 780 (9.9%) | 63977 (9.9%) | 64757 (9.9%) |
| 2004 | 645 (8.2%) | 64256 (10.0%) | 64901 (9.9%) |
| 2005 | 548 (7.0%) | 63973 (9.9%) | 64521 (9.9%) |
| 2006 | 446 (5.7%) | 64807 (10.0%) | 65253 (10.0%) |
| 2007 | 327 (4.2%) | 64519 (10.0%) | 64846 (9.9%) |
| Total number of level 3 ICD-10 diagnoses in the 24-month period before pregnancy, mean (SD) | 9.9 (8.2) | 9.3 (7.9) | 9.3 (7.9) |

### Associations between maternal diagnoses and offspring ASD

There were 331 distinct ICD-10 diagnoses recorded in women during the exposure window. Following the exclusion of rare diagnoses (**Figure S1**), the number of non-chronic and chronic diagnostic categories included in the analyses were 135 and 41, respectively.

#### Phase 1 analyses

After controlling for the study covariates, 40 of the 135 non-chronic maternal diagnoses were nominally significantly associated with ASD (P-value <0.05), 22 of which remained significant after adjusting for multiple testing (q-value <0.05) (**Table 2**), including non-infective gastroenteritis and colitis (HR= 1.87, 95% CI=1.22-2.88), diabetes mellitus in pregnancy (HR=1.26, 95% CI=1.06-1.48) and fracture of skull and facial bones (HR=1.81, 95% CI=1.23-2.66).

**Table 2.** Associations between maternal non-chronic ICD-10 level 3 diagnostic categories and offspring ASD

|  | ICD-10 level 3 | Diagnosis/Medical code | Univariate adjusted model<sup>a</sup> | Multivariable model<sup>b</sup> | Exposed |  |
| --- | --- | --- | --- | --- | --- | --- |
|  |  |  | HR (95%CI) | HR (95%CI) | ASD | control |
| **Non-chronic** | K52 | Other and unspecified noninfective gastroenteritis and colitis | 1.87 [1.22, 2.88]* | 1.67 [1.08, 2.57]† | 21 | 834 |
|  | M62 | Other disorders of muscle | 1.44 [1.17, 1.77]** | 1.29 [1.05, 1.59]† | 95 | 4528 |
|  | N30 | Cystitis | 1.33 [1.08, 1.66]* | 1.20 [0.97, 1.49] | 89 | 4823 |
|  | O24 | Diabetes mellitus in pregnancy, childbirth, and the puerperium | 1.26 [1.06, 1.48]* | 1.28 [1.08, 1.51]†† | 150 | 9609 |
|  | O30 | Multiple gestation | 0.79 [0.69, 0.90]** | 0.81 [0.67, 0.97]† | 260 | 24683 |
|  | O36 | Maternal care for other known or suspected fetal problems | 1.16 [1.05, 1.27]* | 1.17 [1.06, 1.29]†† | 460 | 32492 |
|  | O47 | False labor | 1.22 [1.11, 1.35]** | 1.20 [1.09, 1.33]††† | 450 | 28277 |
|  | O70 | Perineal laceration during delivery | 0.81 [0.75, 0.88]*** | 0.90 [0.83, 0.98]† | 700 | 64428 |
|  | O80 | Encounter for full-term uncomplicated delivery | 0.89 [0.84, 0.94]*** | 1.01 [0.94, 1.09] | 1880 | 152052 |
|  | O84 | Multiple births | 0.74 [0.62, 0.89]* | 0.96 [0.76, 1.23] | 145 | 13468 |
|  | O99 | Other maternal diseases classifiable elsewhere but complicating pregnancy, childbirth and the puerperium | 1.23 [1.11, 1.36]*** | 1.20 [1.09, 1.33]††† | 518 | 43137 |
|  | R10 | Abdominal and pelvic pain | 1.21 [1.08, 1.35]** | 1.11 [0.99, 1.24] | 362 | 22958 |
|  | S02 | Fracture of skull and facial bones | 1.81 [1.23, 2.66]* | 1.61 [1.09, 2.37]† | 26 | 1023 |
|  | S40 | Superficial injury of shoulder and upper arm | 1.51 [1.11, 2.05]* | 1.35 [0.99, 1.83] | 41 | 2061 |
|  | S60 | Superficial injury of wrist, hand and fingers | 1.33 [1.13, 1.56]** | 1.21 [1.03, 1.43]† | 147 | 8263 |
|  | T14 | Injury of unspecified body region | 1.68 [1.31, 2.16]*** | 1.54 [1.19, 1.98]††† | 61 | 2638 |
|  | Z00 | Encounter for general examination without complaint, suspected or reported diagnosis | 0.59 [0.43, 0.82]* | 0.59 [0.43, 0.82]†† | 37 | 4844 |
|  | Z29 | Need for prophylactic measures | 0.80 [0.67, 0.94]* | 0.86 [0.73, 1.02] | 142 | 13519 |
|  | Z32 | Encounter for pregnancy test and childbirth and childcare instruction | 1.09 [1.04, 1.15]** | 1.08 [1.02, 1.13]†† | 2379 | 179850 |
|  | Z36 | Encounter for antenatal screening of mother | 1.07 [1.02, 1.12]* | 1.07 [1.02, 1.12]†† | 2872 | 240698 |
|  | Z39 | Encounter for maternal postpartum care and examination | 0.87 [0.82, 0.92]*** | 1.02 [0.95, 1.09] | 2366 | 183389 |
|  | Z71 | Persons encountering health services for other counseling and medical advice, not elsewhere classified | 0.79 [0.67, 0.92]* | 0.78 [0.67, 0.91]†† | 163 | 20384 |

Note: Statistically significant positive associations from the multivariable model are highlighted in blue and negative associations are highlighted in yellow.
<sup>a</sup> models adjusted for maternal age at childbirth, total number of diagnoses in 24 months preceding childbirth, and year of childbirth
<sup>b</sup> models adjusted for maternal age at childbirth, total number of diagnoses in 24 months preceding childbirth, year of childbirth, and all the ICD-10 diagnostic categories presented in tables 2 and 3
Q: values are p values adjusted for false discovery rate
\*: Q-value <0.05; \*\*: Q-value <0.01; \*\*\*: Q-value <0.001
†: P-value <0.05; ††: P-value <0.01; †††: P-value <0.001

Similarly, 14 of the 41 chronic maternal diagnoses were nominally significantly associated with offspring ASD, and 11 remained significant after adjusting for multiple testing (q-value <0.05) (**Table S2** and **Table 3**), including recurrent major depressive disorder (HR=2.45, 95% CI=1.86, 3.21), reaction to severe stress, adjustment disorders (HR= 1.50, 95% CI=1.27-1.78), and irritable bowel syndrome (HR =1.53, 95% CI=1.15-2.04).

**Table 3.** Associations between maternal chronic ICD-10 level 3 diagnostic categories and offspring ASD

|  | ICD-10 level 3 | Diagnosis/Medical code | Univariate adjusted model<sup>a</sup> | Multivariable model<sup>b</sup> | Exposed |  |
| --- | --- | --- | --- | --- | --- | --- |
|  |  |  | HR (95%CI) | HR (95%CI) | ASD | control |
| **Chronic** | F10 | Alcohol related disorders | 1.54 [1.21, 1.97]** | 1.22 [0.95, 1.57] | 66 | 3203 |
|  | F32 | Major depressive disorder, single episode | 1.53 [1.20, 1.96]** | 1.04 [0.79, 1.38] | 66 | 3733 |
|  | F33 | Major depressive disorder, recurrent | 2.45 [1.86, 3.21]*** | 1.89 [1.39, 2.56]††† | 52 | 1928 |
|  | F41 | Other anxiety disorders | 1.69 [1.27, 2.25]** | 1.33 [0.99, 1.78] | 48 | 2345 |
|  | F43 | Reaction to severe stress, and adjustment disorders | 1.50 [1.27, 1.78]*** | 1.21 [1.01, 1.45]† | 142 | 7552 |
|  | F60 | Specific personality disorders | 1.77 [1.43, 2.18]*** | 1.26 [0.99, 1.60] | 92 | 4026 |
|  | F99 | Mental disorder, not otherwise specified | 1.76 [1.20, 2.60]* | 1.25 [0.83, 1.88] | 26 | 1200 |
|  | G40 | Epilepsy and recurrent seizures | 1.64 [1.30, 2.08]*** | 1.49 [1.17, 1.89]†† | 72 | 3331 |
|  | K07 | Dentofacial anomalies | 1.72 [1.19, 2.50]* | 1.62 [1.12, 2.35]† | 28 | 1428 |
|  | K58 | Irritable bowel syndrome | 1.53 [1.15, 2.04]* | 1.41 [1.05, 1.89]† | 48 | 2574 |
|  | UT0 | Non-smoker mother | 0.74 [0.69, 0.78]*** | 0.79 [0.74, 0.85]††† | 2257 | 198111 |

Note: Statistically significant positive associations from the multivariable model are highlighted in blue and negative associations are highlighted in yellow.
<sup>a</sup> models adjusted for maternal age at childbirth, total number of diagnoses in 24 months preceding childbirth, and year of childbirth
<sup>b</sup> models adjusted for maternal age at childbirth, total number of diagnoses in 24 months preceding childbirth, year of childbirth, and all the ICD-10 diagnostic categories presented in tables 2 and 3
\*: Q-value <0.05; \*\*: Q-value <0.01; \*\*\*: Q-value <0.001
†: P-value <0.05; ††: P-value <0.01; †††: P-value <0.001

In the multivariable model, 15 of 22 non-chronic (**Table 2**) and 6 of 11 chronic (**Table 3**) maternal diagnoses were statistically significantly (p-value < 0.05) associated with offspring ASD. Most maternal non-chronic diagnoses significantly associated with ASD were conditions related to injuries or pregnancy complications. Most of the statistically significant maternal chronic diagnoses associated with increased risk for ASD were mental health and digestive system disorders.

#### Phase 2 analysis

##### Timing of maternal diagnosis

Adding more distal diagnoses of non-chronic conditions to the multivariable model did not change the direction and magnitude of the association between the maternal non-chronic diagnoses occurring within 24 months before childbirth and the risk of ASD (**Figure 1** and **Table S3**). Furthermore, most of the non-chronic maternal diagnoses recorded in the 24-48 months before childbirth were not associated with the risk of ASD; the nominally significant associations observed for several diagnoses were not statistically significant after correction for multiple testing.

**Figure 1.**
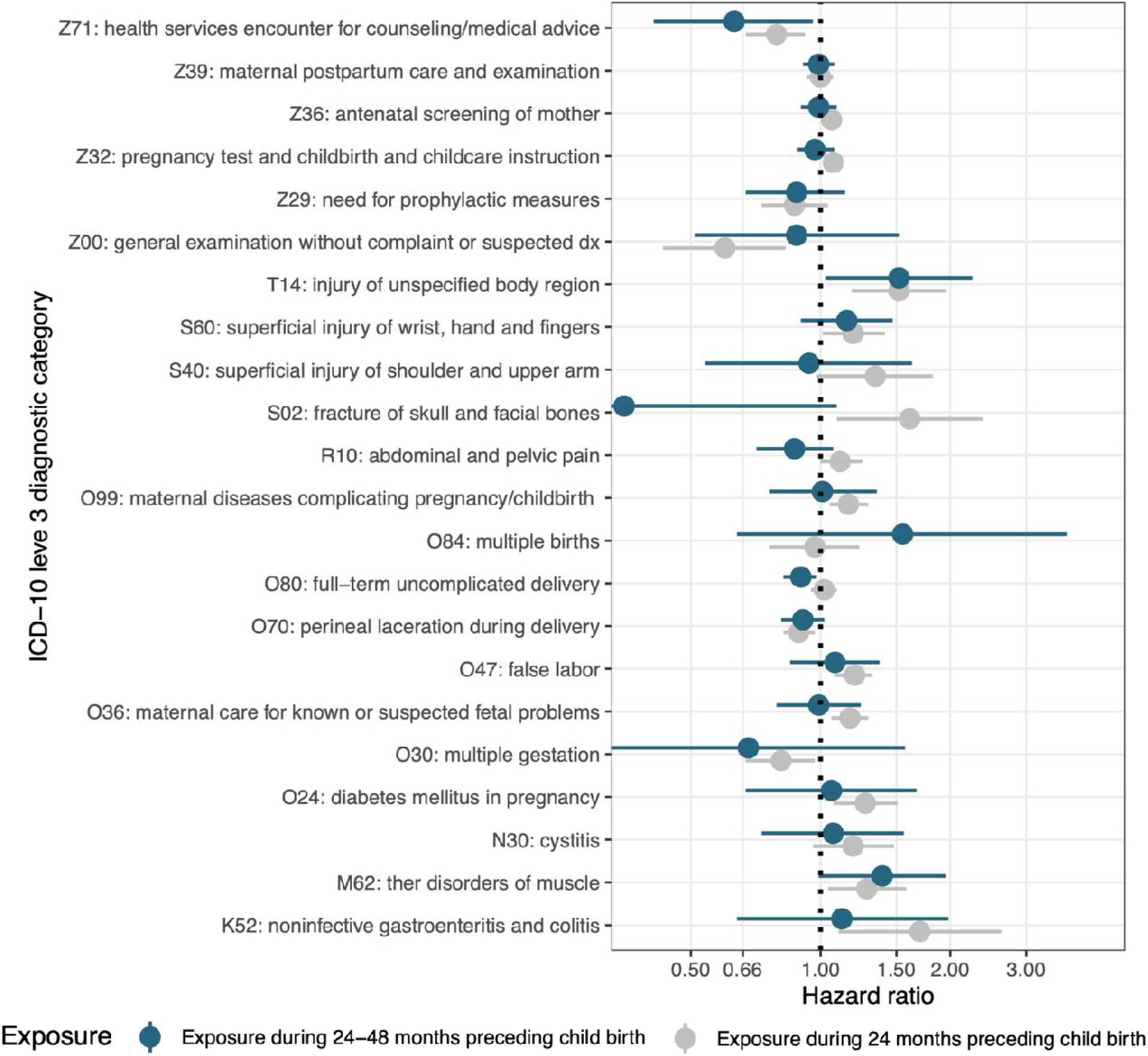
Associations between ICD-10 level 3 maternal diagnoses and offspring ASD by the timing of exposure Note: models adjusted for maternal age at childbirth, total number of diagnoses in 24 months preceding childbirth, year of childbirth, and all the ICD-10 diagnostic categories presented in this figure and chronic conditions: ICD-10: F10, F32, F33, F41, F43, F60, F99, G40, K07, K58

##### Sibling analysis

The sample for the sibling analysis included 418,008 children. The results from the sibling analysis (Figure 2 and Table S4), when compared with the multivariable analysis conducted on the same sample (Figure 2), differed across the maternal diagnoses. For certain associations, such as diagnoses related to maternal injury, the point estimates were very similar, suggesting limited confounding by time-invariant, family-level factors. The attenuation or strengthening of the associations for other associations, for example perinatal complications, were suggestive of possible confounding by family level shared factors that were uncontrolled in the multivariable analysis. It is worth noting that despite the large sample size, estimates from the sibling analysis generally had wide confidence intervals hindering clear conclusions.

**Figure 2.**
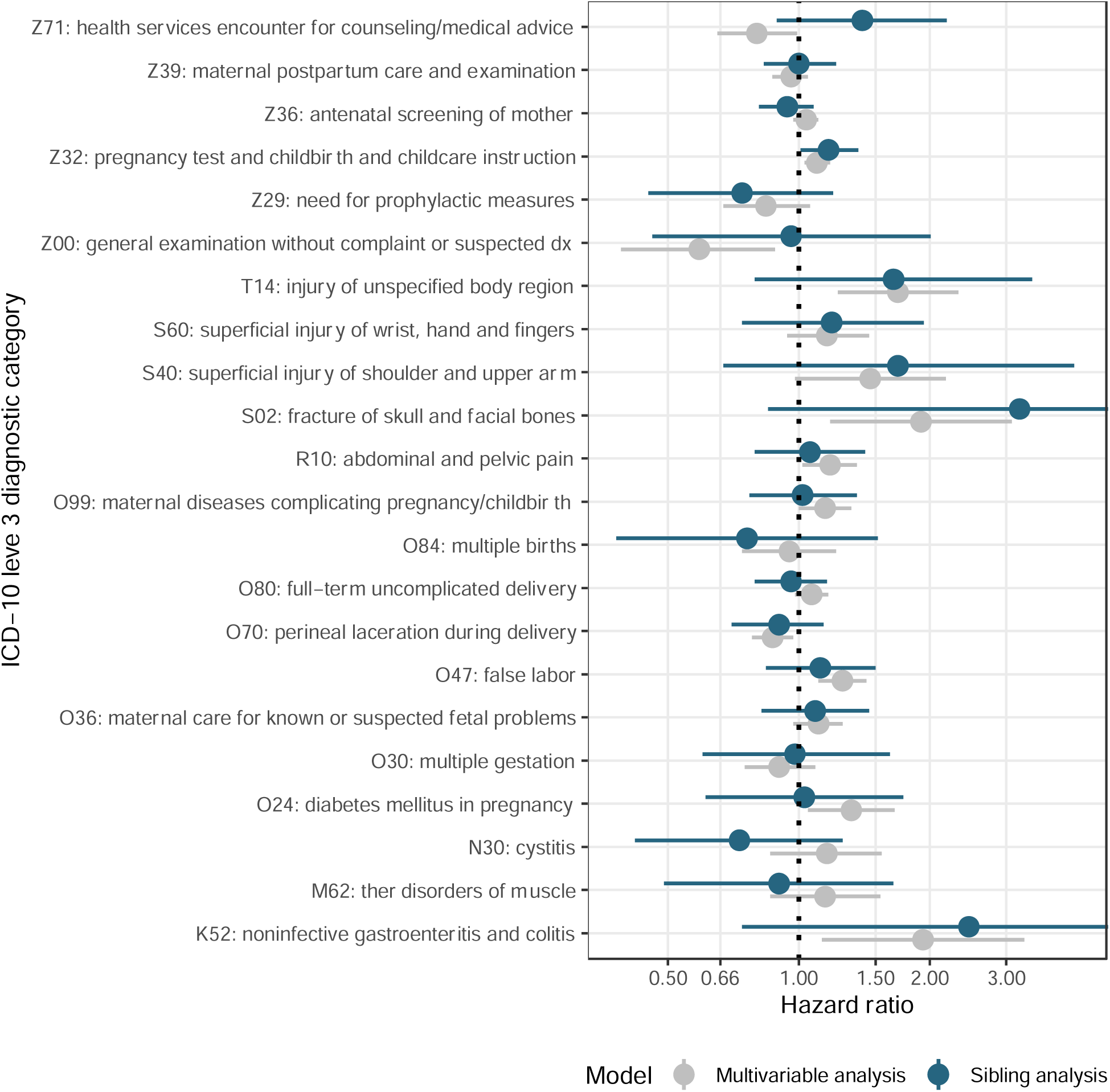
Associations between ICD-10 level 3 maternal diagnoses and offspring ASD from multivariable and sibling analyses Note: models adjusted for maternal age at childbirth, total number of diagnoses in 24 months preceding childbirth, year of childbirth, and all the ICD-10 diagnostic categories presented in this figure and chronic conditions: ICD-10: F10, F32, F33, F41, F43, F60, F99, G40, K07, K58 The multivariable and sibling analyses presented in this figure were restricted to individuals with at least one sibling in the sample (including 418,008 of the original 653,580 mother child dyads). The potential differences between the results from the multivariable analysis in the subsample of siblings only and the full sample could be attributable to potential differences in sample composition and sample size. Comments for fig 2 like fig 1

## DISCUSSION

In this population-based study of more than 600,000 pregnancies we evaluated the associations between the range of maternal diagnoses around pregnancy with the risk of ASD in the offspring. Of 331 maternal diagnoses assessed, we observed 15 non-chronic and 6 chronic that were associated with offspring ASD after adjusting for child’s sex and year of birth, maternal age at childbirth, and the total number of maternal diagnoses. Our results highlight the importance of maternal health in pregnancy in ASD risk and offer novel hypotheses regarding the etiology of ASD – necessitating further dissection of the mechanisms underlying the observed associations.

Most of the non-chronic maternal diagnoses associated with the risk of ASD in offspring concerned injury (unrelated to pregnancy). Head and upper body injuries, and injuries to the unspecified body region were associated with an increased risk of ASD. Replication of the results in sibling analysis along with the absence of associations between injuries occurring long before pregnancy were suggestive that the observed associations are robust to potential familial confounders. The studies of maternal physical trauma and risk of ASD remain scarce^22,23^, with limited evidence for the effect of maternal trauma in pregnancy on the risk of autism^22^.

A potential mechanism for this associations could be the interference of maternal trauma with neurodevelopment of the fetus. Various studies have pointed to the adverse effects of trauma on fetal and child health outcomes, including neurodevelopment. There is also evidence that traumas, even in the absence of notable maternal distress, could affect the fetus^24^. Another potential mechanism underlying these associations could be the role of immune activation in trauma^25,26^. Mounting evidence suggests that maternal immune activation (MIA) may be a risk factor for neurodevelopmental and behavioral abnormalities in the offspring^27^. Although the majority of studies of inflammation and neurodevelopmental outcomes have mainly focused on infections in pregnancy^27^, other conditions associated with maternal acute and chronic inflammation have been linked to neurodevelopmental disorders^28^. Therefore, it is possible that the observed association between maternal injury and ASD is attributable to the MIA.

Additionally, to our knowledge, the current study is the first to report an association between offspring ASD and maternal noninfective gastroenteritis and colitis. The results from timing of maternal diagnosis and sibling analyses suggested that the observed association was not attributable to shared family level factors. We also observed a similar association between ASD and irritable bowel syndrome (IBS) diagnosis in the mother. It is plausible that inflammation and immune system activation in gastroenteritis and colitis as well as IBS could account for the observed associations^28^. The potential impact of these conditions on maternal diet in pregnancy^29,30^ could also be another possible mechanism through which these conditions could affect the risk of ASD in the offspring^31^. Our findings warrant studies further exploring these associations and the potential mechanisms underlying them.

Prior studies have reported associations between pregnancy complications and neurodevelopmental disorders, including ASD^32,33^. We demonstrated that many of these diagnoses are not associated with child’s ASD if they occurred during the 24-48 months before the childbirth (i.e., in a previous pregnancy), suggesting the results were unlikely to arise due to familial confounding. Nevertheless, the estimates from the sibling analysis had wide confidence intervals, limiting the inference regarding familial confounding.

Maternal mental health conditions, including depression, reaction to severe stress, and adjustment disorders, were the main group of chronic disorders associated with increased risk of offspring ASD. Multiple epidemiological studies have reported on an association between maternal stress, depression, and anxiety, and offspring neurodevelopmental outcomes^34^, including ASD^35^. Our results show that these associations are not due to confounding by other diagnoses that were found to be associated with ASD as well as a number of maternal factors potentially associated with mental illness, including age at conceiving the child^36^ and healthcare utilization.

This is the first comprehensive assessment of the range of maternal diagnoses around pregnancy for their associations with ASD in the offspring. The systematic nature of our analysis allowed for a direct comparison of the estimated associations between different maternal diagnoses and ASD in the offspring. Additional strengths of this study include classification of diagnoses into chronic and non-chronic categories, interrogating the effects by exposure timing, and accounting for familial confounding in the sibling analysis.

The study also has several limitations. Firstly, we did not account for maternal medication use. If a diagnosis arises due to medication (e.g., as its side effect), rather than being the reason for medication use, it could result in a spurious association if the same medication is also associated with the risk of offspring ASD. We attempted to minimize the potential of such bias by concurrently including all diagnoses statistically significantly associated with ASD in the multivariable model. Secondly, we did not have information about several sociodemographic factors, which limited our ability to control for potential effects of these characteristics (e.g., education, ethnicity, immigration status, and household income). Finally, ascertaining maternal diagnosis using at least 1 record of specific diagnostic codes offered high sensitivity, nevertheless, it could have resulted in lower specificity (e.g., due to administrative errors, or misdiagnosis), affecting the estimated associations.

In conclusion, the current study harnessed multiple analytic approaches to provide insights into the associations between maternal diagnoses during pregnancy and the risk of ASD in offspring. We identified new associations between maternal chronic and non-chronic diagnoses, replicated some associations established previously, and evaluated the role of familial confounding in these observations. The current study findings draw attention to the importance of maternal health, specifically around pregnancy, in the risk of offspring ASD. Study findings highlight avenues for further exploration of etiologic mechanisms in ASD, and suggest that careful monitoring of early child development may be warranted for children of mothers with a high burden of certain diagnoses around the time of pregnancy.

## Data Availability

This project uses the Danish national register data. The Danish Scientific Ethical Committee system, as well as all relevant register authorities, including the Danish Data Protection Agency, Statistics Denmark, and the Danish Health Data Authority, approved access to these data under strict conditions regarding access and data export. Under these conditions there are no provisions for exporting individual level data, all or in part, to another institution, in or outside of Denmark. Individual-level data is available to verified and approved researchers, but can never be exported outside of the Danish servers.

## ACKNOWLEDGMENTS

We would like to acknowledge the generous support of the Seaver Foundation. This work was supported in part by a grant from the National Institute of Mental Health (Dr. Janecka, Dr. Reichenberg, Dr. Sandin [grant number MH124817], Dr. Khachadourian [Award number T32-MH122394]). The content is solely the responsibility of the authors and does not necessarily represent the official views of the funding agencies or the authors’ employers.

## SUPPLEMENTAL MATERIALS

**Figure S1.**
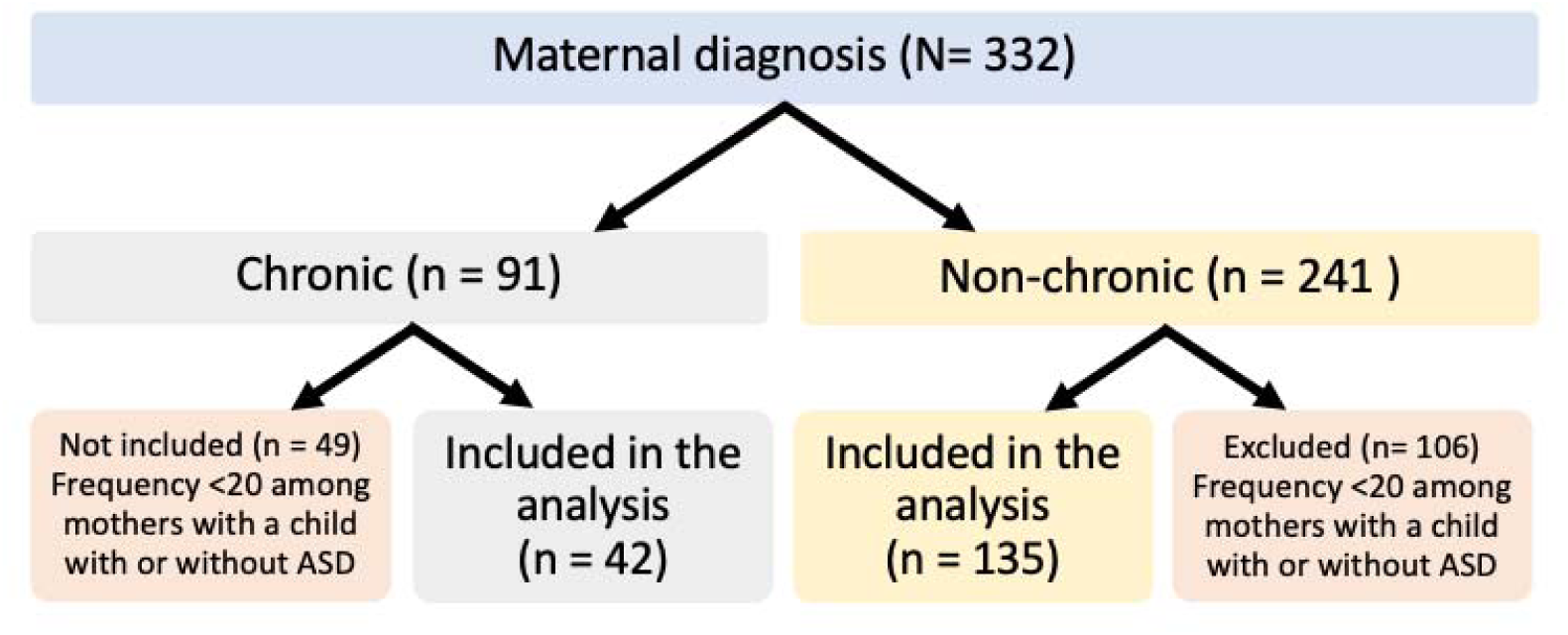
Flowchart of maternal diagnosis around pregnancy

**Table S1.**
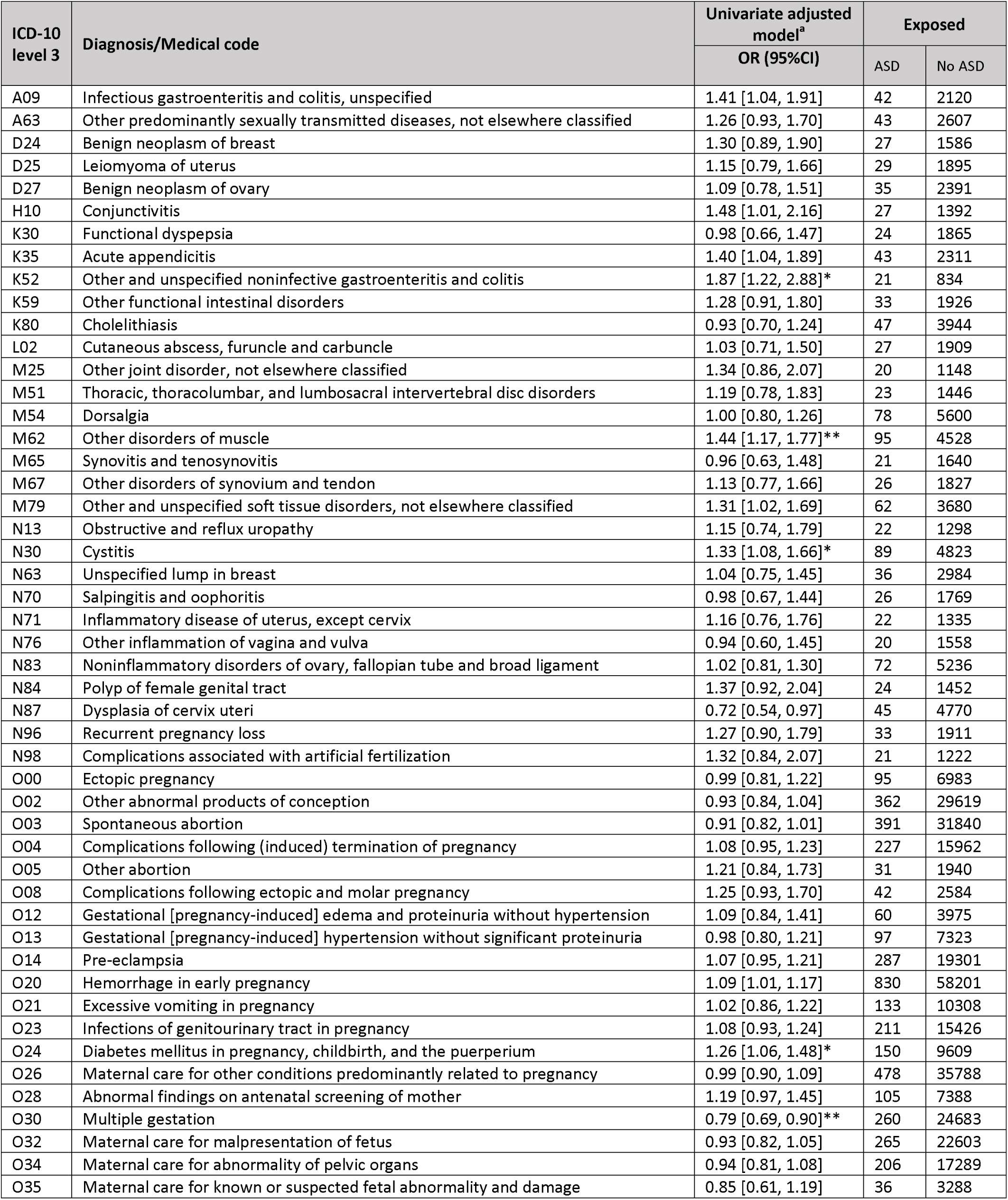
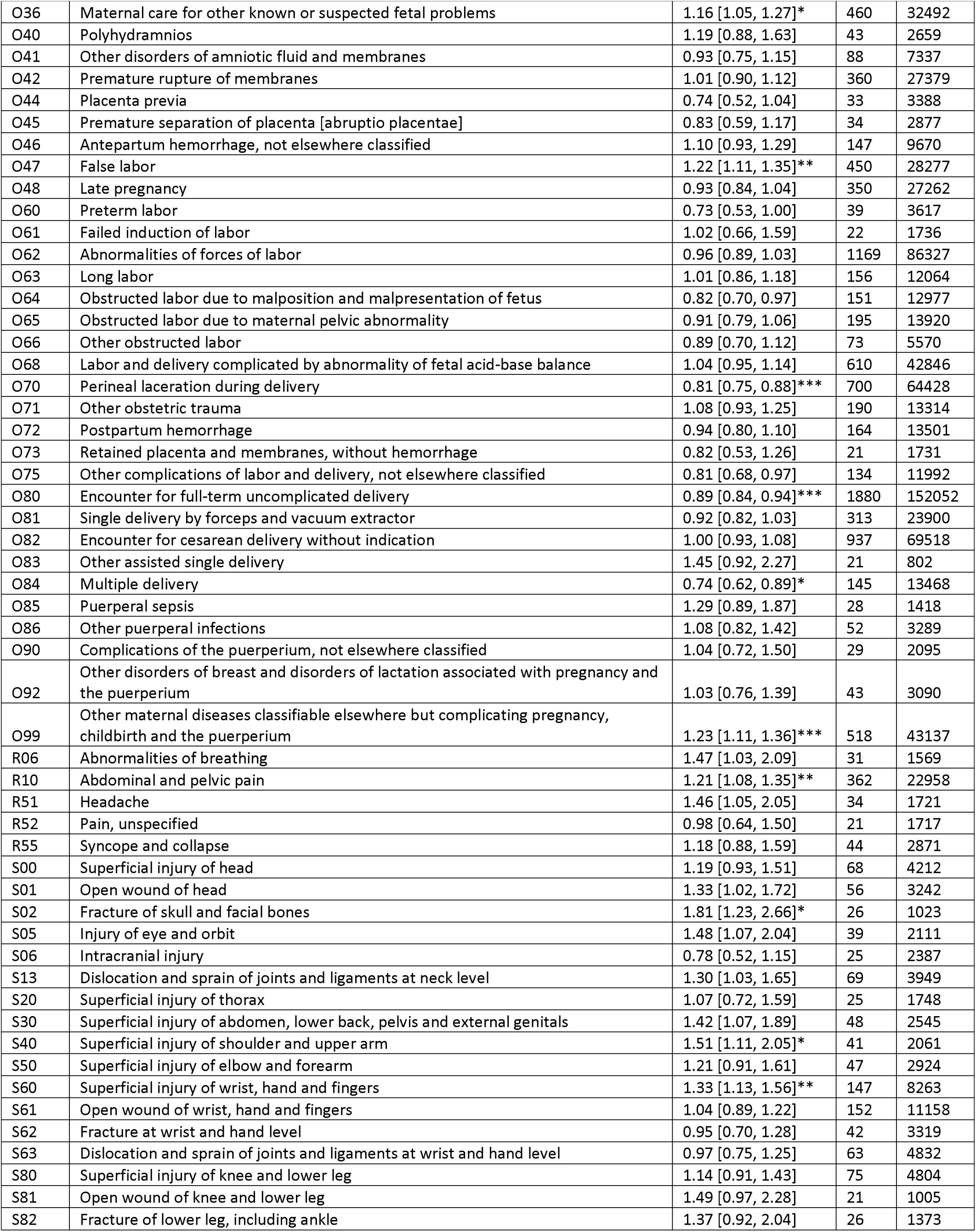
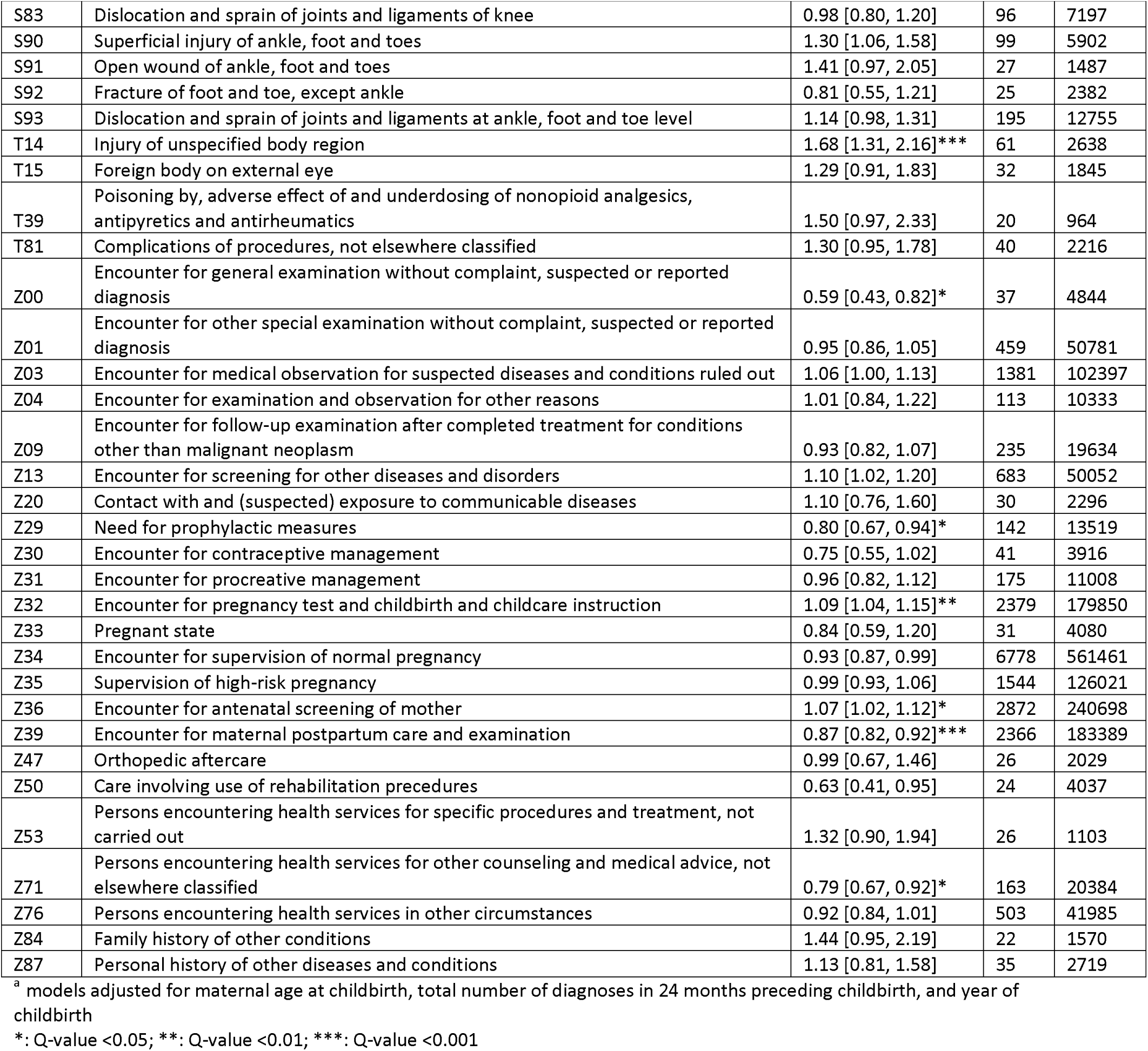
Associations between maternal ICD-10 level 3 non-chronic diagnostic categories and offspring ASD in univariate models, adjusted for covariates.

| ICD-10 level 3 | Diagnosis/Medical code | Univariate adjusted model<sup>a</sup><br>OR (95%CI) | Exposed |  |
| --- | --- | --- | --- | --- |
|  |  |  | ASD | No ASD |
| A09 | Infectious gastroenteritis and colitis, unspecified | 1.41 [1.04, 1.91] | 42 | 2120 |
| A63 | Other predominantly sexually transmitted diseases, not elsewhere classified | 1.26 [0.93, 1.70] | 43 | 2607 |
| D24 | Benign neoplasm of breast | 1.30 [0.89, 1.90] | 27 | 1586 |
| D25 | Leiomyoma of uterus | 1.15 [0.79, 1.66] | 29 | 1895 |
| D27 | Benign neoplasm of ovary | 1.09 [0.78, 1.51] | 35 | 2391 |
| H10 | Conjunctivitis | 1.48 [1.01, 2.16] | 27 | 1392 |
| K30 | Functional dyspepsia | 0.98 [0.66, 1.47] | 24 | 1865 |
| K35 | Acute appendicitis | 1.40 [1.04, 1.89] | 43 | 2311 |
| K52 | Other and unspecified noninfective gastroenteritis and colitis | 1.87 [1.22, 2.88]* | 21 | 834 |
| K59 | Other functional intestinal disorders | 1.28 [0.91, 1.80] | 33 | 1926 |
| K80 | Cholelithiasis | 0.93 [0.70, 1.24] | 47 | 3944 |
| L02 | Cutaneous abscess, furuncle and carbuncle | 1.03 [0.71, 1.50] | 27 | 1909 |
| M25 | Other joint disorder, not elsewhere classified | 1.34 [0.86, 2.07] | 20 | 1148 |
| M51 | Thoracic, thoracolumbar, and lumbosacral intervertebral disc disorders | 1.19 [0.78, 1.83] | 23 | 1446 |
| M54 | Dorsalgia | 1.00 [0.80, 1.26] | 78 | 5600 |
| M62 | Other disorders of muscle | 1.44 [1.17, 1.77]** | 95 | 4528 |
| M65 | Synovitis and tenosynovitis | 0.96 [0.63, 1.48] | 21 | 1640 |
| M67 | Other disorders of synovium and tendon | 1.13 [0.77, 1.66] | 26 | 1827 |
| M79 | Other and unspecified soft tissue disorders, not elsewhere classified | 1.31 [1.02, 1.69] | 62 | 3680 |
| N13 | Obstructive and reflux uropathy | 1.15 [0.74, 1.79] | 22 | 1298 |
| N30 | Cystitis | 1.33 [1.08, 1.66]* | 89 | 4823 |
| N63 | Unspecified lump in breast | 1.04 [0.75, 1.45] | 36 | 2984 |
| N70 | Salpingitis and oophoritis | 0.98 [0.67, 1.44] | 26 | 1769 |
| N71 | Inflammatory disease of uterus, except cervix | 1.16 [0.76, 1.76] | 22 | 1335 |
| N76 | Other inflammation of vagina and vulva | 0.94 [0.60, 1.45] | 20 | 1558 |
| N83 | Noninflammatory disorders of ovary, fallopian tube and broad ligament | 1.02 [0.81, 1.30] | 72 | 5236 |
| N84 | Polyp of female genital tract | 1.37 [0.92, 2.04] | 24 | 1452 |
| N87 | Dysplasia of cervix uteri | 0.72 [0.54, 0.97] | 45 | 4770 |
| N96 | Recurrent pregnancy loss | 1.27 [0.90, 1.79] | 33 | 1911 |
| N98 | Complications associated with artificial fertilization | 1.32 [0.84, 2.07] | 21 | 1222 |
| O00 | Ectopic pregnancy | 0.99 [0.81, 1.22] | 95 | 6983 |
| O02 | Other abnormal products of conception | 0.93 [0.84, 1.04] | 362 | 29619 |
| O03 | Spontaneous abortion | 0.91 [0.82, 1.01] | 391 | 31840 |
| O04 | Complications following (induced) termination of pregnancy | 1.08 [0.95, 1.23] | 227 | 15962 |
| O05 | Other abortion | 1.21 [0.84, 1.73] | 31 | 1940 |
| O08 | Complications following ectopic and molar pregnancy | 1.25 [0.93, 1.70] | 42 | 2584 |
| O12 | Gestational [pregnancy-induced] edema and proteinuria without hypertension | 1.09 [0.84, 1.41] | 60 | 3975 |
| O13 | Gestational [pregnancy-induced] hypertension without significant proteinuria | 0.98 [0.80, 1.21] | 97 | 7323 |
| O14 | Pre-eclampsia | 1.07 [0.95, 1.21] | 287 | 19301 |
| O20 | Hemorrhage in early pregnancy | 1.09 [1.01, 1.17] | 830 | 58201 |
| O21 | Excessive vomiting in pregnancy | 1.02 [0.86, 1.22] | 133 | 10308 |
| O23 | Infections of genitourinary tract in pregnancy | 1.08 [0.93, 1.24] | 211 | 15426 |
| O24 | Diabetes mellitus in pregnancy, childbirth, and the puerperium | 1.26 [1.06, 1.48]* | 150 | 9609 |
| O26 | Maternal care for other conditions predominantly related to pregnancy | 0.99 [0.90, 1.09] | 478 | 35788 |
| O28 | Abnormal findings on antenatal screening of mother | 1.19 [0.97, 1.45] | 105 | 7388 |
| O30 | Multiple gestation | 0.79 [0.69, 0.90]** | 260 | 24683 |
| O32 | Maternal care for malpresentation of fetus | 0.93 [0.82, 1.05] | 265 | 22603 |
| O34 | Maternal care for abnormality of pelvic organs | 0.94 [0.81, 1.08] | 206 | 17289 |
| O35 | Maternal care for known or suspected fetal abnormality and damage | 0.85 [0.61, 1.19] | 36 | 3288 |
| O36 | Maternal care for other known or suspected fetal problems | 1.16 [1.05, 1.27]* | 460 | 32492 |
| O40 | Polyhydramnios | 1.19 [0.88, 1.63] | 43 | 2659 |
| O41 | Other disorders of amniotic fluid and membranes | 0.93 [0.75, 1.15] | 88 | 7337 |
| O42 | Premature rupture of membranes | 1.01 [0.90, 1.12] | 360 | 27379 |
| O44 | Placenta previa | 0.74 [0.52, 1.04] | 33 | 3388 |
| O45 | Premature separation of placenta [abruptio placentae] | 0.83 [0.59, 1.17] | 34 | 2877 |
| O46 | Antepartum hemorrhage, not elsewhere classified | 1.10 [0.93, 1.29] | 147 | 9670 |
| O47 | False labor | 1.22 [1.11, 1.35]** | 450 | 28277 |
| O48 | Late pregnancy | 0.93 [0.84, 1.04] | 350 | 27262 |
| O60 | Preterm labor | 0.73 [0.53, 1.00] | 39 | 3617 |
| O61 | Failed induction of labor | 1.02 [0.66, 1.59] | 22 | 1736 |
| O62 | Abnormalities of forces of labor | 0.96 [0.89, 1.03] | 1169 | 86327 |
| O63 | Long labor | 1.01 [0.86, 1.18] | 156 | 12064 |
| O64 | Obstructed labor due to malposition and malpresentation of fetus | 0.82 [0.70, 0.97] | 151 | 12977 |
| O65 | Obstructed labor due to maternal pelvic abnormality | 0.91 [0.79, 1.06] | 195 | 13920 |
| O66 | Other obstructed labor | 0.89 [0.70, 1.12] | 73 | 5570 |
| O68 | Labor and delivery complicated by abnormality of fetal acid-base balance | 1.04 [0.95, 1.14] | 610 | 42846 |
| O70 | Perineal laceration during delivery | 0.81 [0.75, 0.88]*** | 700 | 64428 |
| O71 | Other obstetric trauma | 1.08 [0.93, 1.25] | 190 | 13314 |
| O72 | Postpartum hemorrhage | 0.94 [0.80, 1.10] | 164 | 13501 |
| O73 | Retained placenta and membranes, without hemorrhage | 0.82 [0.53, 1.26] | 21 | 1731 |
| O75 | Other complications of labor and delivery, not elsewhere classified | 0.81 [0.68, 0.97] | 134 | 11992 |
| O80 | Encounter for full-term uncomplicated delivery | 0.89 [0.84, 0.94]*** | 1880 | 152052 |
| O81 | Single delivery by forceps and vacuum extractor | 0.92 [0.82, 1.03] | 313 | 23900 |
| O82 | Encounter for cesarean delivery without indication | 1.00 [0.93, 1.08] | 937 | 69518 |
| O83 | Other assisted single delivery | 1.45 [0.92, 2.27] | 21 | 802 |
| O84 | Multiple delivery | 0.74 [0.62, 0.89]* | 145 | 13468 |
| O85 | Puerperal sepsis | 1.29 [0.89, 1.87] | 28 | 1418 |
| O86 | Other puerperal infections | 1.08 [0.82, 1.42] | 52 | 3289 |
| O90 | Complications of the puerperium, not elsewhere classified | 1.04 [0.72, 1.50] | 29 | 2095 |
| O92 | Other disorders of breast and disorders of lactation associated with pregnancy and the puerperium | 1.03 [0.76, 1.39] | 43 | 3090 |
| O99 | Other maternal diseases classifiable elsewhere but complicating pregnancy, childbirth and the puerperium | 1.23 [1.11, 1.36]*** | 518 | 43137 |
| R06 | Abnormalities of breathing | 1.47 [1.03, 2.09] | 31 | 1569 |
| R10 | Abdominal and pelvic pain | 1.21 [1.08, 1.35]** | 362 | 22958 |
| R51 | Headache | 1.46 [1.05, 2.05] | 34 | 1721 |
| R52 | Pain, unspecified | 0.98 [0.64, 1.50] | 21 | 1717 |
| R55 | Syncope and collapse | 1.18 [0.88, 1.59] | 44 | 2871 |
| S00 | Superficial injury of head | 1.19 [0.93, 1.51] | 68 | 4212 |
| S01 | Open wound of head | 1.33 [1.02, 1.72] | 56 | 3242 |
| S02 | Fracture of skull and facial bones | 1.81 [1.23, 2.66]* | 26 | 1023 |
| S05 | Injury of eye and orbit | 1.48 [1.07, 2.04] | 39 | 2111 |
| S06 | Intracranial injury | 0.78 [0.52, 1.15] | 25 | 2387 |
| S13 | Dislocation and sprain of joints and ligaments at neck level | 1.30 [1.03, 1.65] | 69 | 3949 |
| S20 | Superficial injury of thorax | 1.07 [0.72, 1.59] | 25 | 1748 |
| S30 | Superficial injury of abdomen, lower back, pelvis and external genitals | 1.42 [1.07, 1.89] | 48 | 2545 |
| S40 | Superficial injury of shoulder and upper arm | 1.51 [1.11, 2.05]* | 41 | 2061 |
| S50 | Superficial injury of elbow and forearm | 1.21 [0.91, 1.61] | 47 | 2924 |
| S60 | Superficial injury of wrist, hand and fingers | 1.33 [1.13, 1.56]** | 147 | 8263 |
| S61 | Open wound of wrist, hand and fingers | 1.04 [0.89, 1.22] | 152 | 11158 |
| S62 | Fracture at wrist and hand level | 0.95 [0.70, 1.28] | 42 | 3319 |
| S63 | Dislocation and sprain of joints and ligaments at wrist and hand level | 0.97 [0.75, 1.25] | 63 | 4832 |
| S80 | Superficial injury of knee and lower leg | 1.14 [0.91, 1.43] | 75 | 4804 |
| S81 | Open wound of knee and lower leg | 1.49 [0.97, 2.28] | 21 | 1005 |
| S82 | Fracture of lower leg, including ankle | 1.37 [0.92, 2.04] | 26 | 1373 |
| S83 | Dislocation and sprain of joints and ligaments of knee | 0.98 [0.80, 1.20] | 96 | 7197 |
| S90 | Superficial injury of ankle, foot and toes | 1.30 [1.06, 1.58] | 99 | 5902 |
| S91 | Open wound of ankle, foot and toes | 1.41 [0.97, 2.05] | 27 | 1487 |
| S92 | Fracture of foot and toe, except ankle | 0.81 [0.55, 1.21] | 25 | 2382 |
| S93 | Dislocation and sprain of joints and ligaments at ankle, foot and toe level | 1.14 [0.98, 1.31] | 195 | 12755 |
| T14 | Injury of unspecified body region | 1.68 [1.31, 2.16]*** | 61 | 2638 |
| T15 | Foreign body on external eye | 1.29 [0.91, 1.83] | 32 | 1845 |
| T39 | Poisoning by, adverse effect of and underdosing of nonopioid analgesics, antipyretics and antirheumatics | 1.50 [0.97, 2.33] | 20 | 964 |
| T81 | Complications of procedures, not elsewhere classified | 1.30 [0.95, 1.78] | 40 | 2216 |
| Z00 | Encounter for general examination without complaint, suspected or reported diagnosis | 0.59 [0.43, 0.82]* | 37 | 4844 |
| Z01 | Encounter for other special examination without complaint, suspected or reported diagnosis | 0.95 [0.86, 1.05] | 459 | 50781 |
| Z03 | Encounter for medical observation for suspected diseases and conditions ruled out | 1.06 [1.00, 1.13] | 1381 | 102397 |
| Z04 | Encounter for examination and observation for other reasons | 1.01 [0.84, 1.22] | 113 | 10333 |
| Z09 | Encounter for follow-up examination after completed treatment for conditions other than malignant neoplasm | 0.93 [0.82, 1.07] | 235 | 19634 |
| Z13 | Encounter for screening for other diseases and disorders | 1.10 [1.02, 1.20] | 683 | 50052 |
| Z20 | Contact with and (suspected) exposure to communicable diseases | 1.10 [0.76, 1.60] | 30 | 2296 |
| Z29 | Need for prophylactic measures | 0.80 [0.67, 0.94]* | 142 | 13519 |
| Z30 | Encounter for contraceptive management | 0.75 [0.55, 1.02] | 41 | 3916 |
| Z31 | Encounter for procreative management | 0.96 [0.82, 1.12] | 175 | 11008 |
| Z32 | Encounter for pregnancy test and childbirth and childcare instruction | 1.09 [1.04, 1.15]** | 2379 | 179850 |
| Z33 | Pregnant state | 0.84 [0.59, 1.20] | 31 | 4080 |
| Z34 | Encounter for supervision of normal pregnancy | 0.93 [0.87, 0.99] | 6778 | 561461 |
| Z35 | Supervision of high-risk pregnancy | 0.99 [0.93, 1.06] | 1544 | 126021 |
| Z36 | Encounter for antenatal screening of mother | 1.07 [1.02, 1.12]* | 2872 | 240698 |
| Z39 | Encounter for maternal postpartum care and examination | 0.87 [0.82, 0.92]*** | 2366 | 183389 |
| Z47 | Orthopedic aftercare | 0.99 [0.67, 1.46] | 26 | 2029 |
| Z50 | Care involving use of rehabilitation procedures | 0.63 [0.41, 0.95] | 24 | 4037 |
| Z53 | Persons encountering health services for specific procedures and treatment, not carried out | 1.32 [0.90, 1.94] | 26 | 1103 |
| Z71 | Persons encountering health services for other counseling and medical advice, not elsewhere classified | 0.79 [0.67, 0.92]* | 163 | 20384 |
| Z76 | Persons encountering health services in other circumstances | 0.92 [0.84, 1.01] | 503 | 41985 |
| Z84 | Family history of other conditions | 1.44 [0.95, 2.19] | 22 | 1570 |
| Z87 | Personal history of other diseases and conditions | 1.13 [0.81, 1.58] | 35 | 2719 |

<sup>a</sup> models adjusted for maternal age at childbirth, total number of diagnoses in 24 months preceding childbirth, and year of childbirth
\*: Q-value <0.05; \*\*: Q-value <0.01; \*\*\*: Q-value <0.001

**Table S2.** Associations between maternal ICD-10 level 3 chronic diagnostic categories and offspring ASD

| ICD-10 level 3 | Diagnosis/Medical code | Univariate adjusted model<sup>a</sup> | Exposed |  |
| --- | --- | --- | --- | --- |
|  |  | OR (95%CI) | ASD | No ASD |
| E03 | Other hypothyroidism | 0.86 [0.60, 1.23] | 30 | 3105 |
| E04 | Other nontoxic goiter | 1.12 [0.80, 1.59] | 36 | 2560 |
| E05 | Thyrototoxicosis [hyperthyroidism] | 1.06 [0.80, 1.42] | 48 | 3930 |
| E10 | Type 1 diabetes mellitus | 0.95 [0.66, 1.39] | 28 | 1956 |
| E28 | Ovarian dysfunction | 1.00 [0.71, 1.42] | 32 | 2939 |
| E66 | Overweight and obesity | 1.07 [0.94, 1.22] | 272 | 29282 |
| F10 | Alcohol related disorders | 1.54 [1.21, 1.97]** | 66 | 3203 |
| F32 | Major depressive disorder, single episode | 1.53 [1.20, 1.96]** | 66 | 3733 |
| F33 | Major depressive disorder, recurrent | 2.45 [1.86, 3.21]*** | 52 | 1928 |
| F40 | Phobic anxiety disorders | 1.10 [0.78, 1.55] | 33 | 2182 |
| F41 | Other anxiety disorders | 1.69 [1.27, 2.25]** | 48 | 2345 |
| F43 | Reaction to severe stress, and adjustment disorders | 1.50 [1.27, 1.78]*** | 142 | 7552 |
| F50 | Eating disorders | 1.54 [1.07, 2.21] | 29 | 1529 |
| F60 | Specific personality disorders | 1.77 [1.43, 2.18]*** | 92 | 4026 |
| F99 | Mental disorder, not otherwise specified | 1.76 [1.20, 2.60]* | 26 | 1200 |
| G40 | Epilepsy and recurrent seizures | 1.64 [1.30, 2.08]*** | 72 | 3331 |
| G43 | Migraine | 1.30 [0.98, 1.72] | 48 | 2899 |
| G56 | Mononeuropathies of upper limb | 0.96 [0.71, 1.31] | 43 | 3990 |
| H91 | Other and unspecified hearing loss | 1.17 [0.78, 1.76] | 23 | 1629 |
| I10 | Essential (primary) hypertension | 1.40 [1.01, 1.96] | 34 | 1977 |
| I47 | Paroxysmal tachycardia | 1.27 [0.87, 1.86] | 27 | 1607 |
| I84 | Haemorrhoids | 0.74 [0.51, 1.05] | 32 | 3606 |
| J30 | Vasomotor and allergic rhinitis | 0.96 [0.67, 1.36] | 31 | 2591 |
| J35 | Chronic diseases of tonsils and adenoids | 1.10 [0.86, 1.41] | 63 | 4482 |
| J45 | Asthma | 1.22 [1.01, 1.49] | 104 | 6924 |
| K07 | Dentofacial anomalies | 1.72 [1.19, 2.50]* | 28 | 1428 |
| K21 | Gastro-esophageal reflux disease | 1.45 [1.00, 2.10] | 28 | 1487 |
| K50 | Crohn's disease [regional enteritis] | 1.05 [0.66, 1.68] | 21 | 1582 |
| K51 | Ulcerative colitis | 1.18 [0.85, 1.65] | 37 | 2541 |
| K58 | Irritable bowel syndrome | 1.53 [1.15, 2.04]* | 48 | 2574 |
| L20 | Atopic dermatitis | 1.41 [0.93, 2.15] | 22 | 1255 |
| M22 | Disorder of patella | 1.24 [0.93, 1.65] | 48 | 2920 |
| M23 | Internal derangement of knee | 0.88 [0.70, 1.12] | 71 | 6652 |
| N60 | Benign mammary dysplasia | 1.11 [0.73, 1.68] | 22 | 1529 |
| N80 | Endometriosis | 0.94 [0.69, 1.28] | 40 | 3541 |
| N92 | Excessive, frequent and irregular menstruation | 0.77 [0.58, 1.02] | 49 | 4831 |
| N94 | Pain and other conditions associated with female genital organs and menstrual cycle | 0.94 [0.69, 1.29] | 42 | 3492 |
| N97 | Female infertility | 0.97 [0.87, 1.07] | 417 | 35423 |
| O10 | Pre-existing hypertension complicating pregnancy, childbirth and the puerperium | 0.95 [0.71, 1.27] | 48 | 3843 |
| O24 | Diabetes mellitus in pregnancy, childbirth, and the puerperium | 1.09 [0.85, 1.40] | 68 | 4431 |
| O99 | Other maternal diseases classifiable elsewhere but complicating pregnancy, childbirth and the puerperium | 0.92 [0.82, 1.04] | 306 | 24080 |
| UT0 | Non-smoker mother | 0.74 [0.69, 0.78]*** | 2257 | 198111 |

<sup>a</sup> models adjusted for maternal age at childbirth, total number of diagnoses in 24 months preceding childbirth, and year of childbirth
\*: Q-value <0.05; \*\*: Q-value <0.01; \*\*\*: Q-value <0.001

**Table S3.**
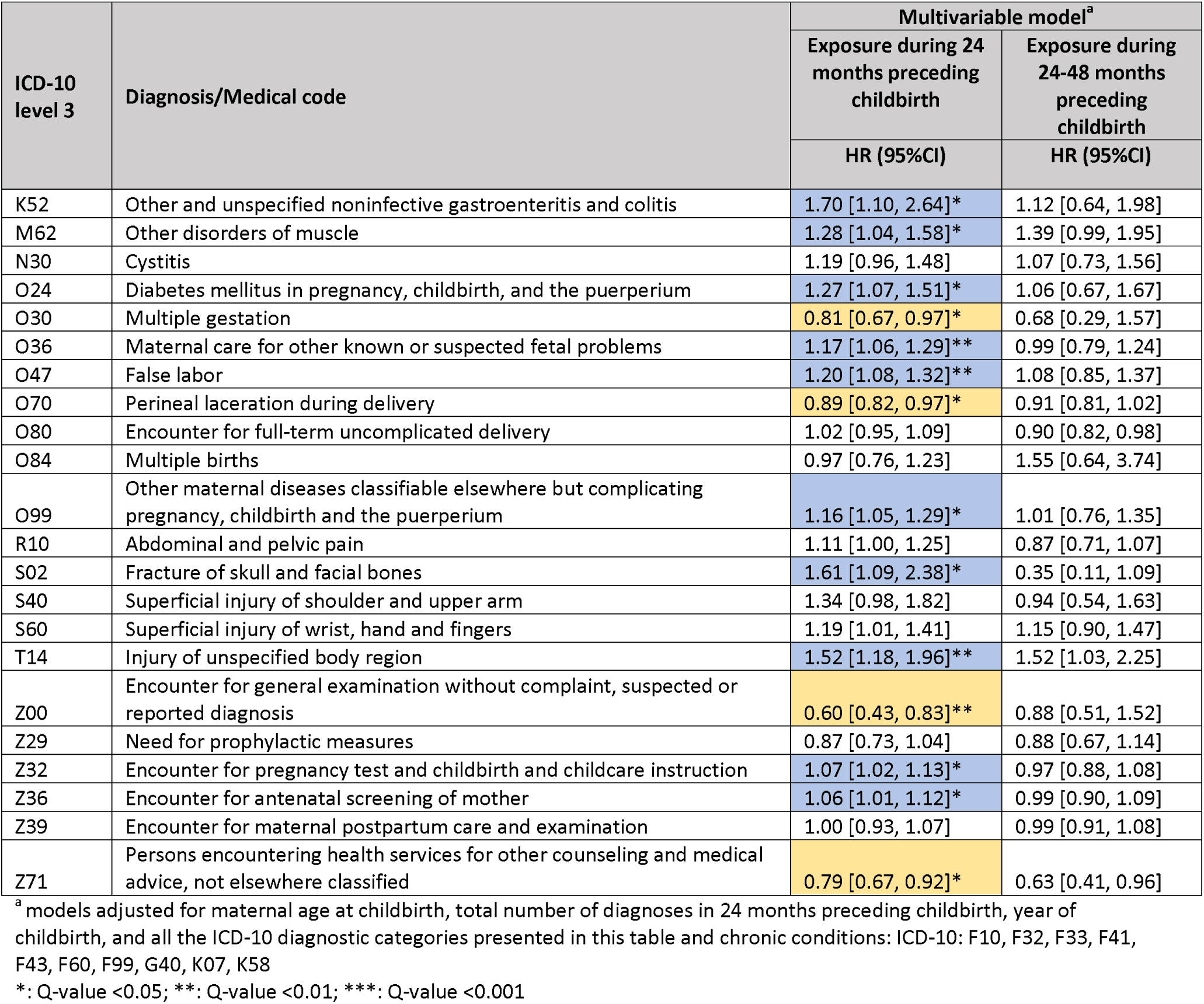
Associations between maternal ICD-10 level 3 non-chronic diagnostic categories and offspring ASD by the time of diagnosis

**Table S4.** Associations between maternal ICD-10 level 3 non-chronic diagnostic categories and offspring ASD from sibling analysis

| ICD-10 level 3 | Diagnosis/Medical code | Multivariable model<sup>a</sup> |
| --- | --- | --- |
|  |  | Exposure during 24 months preceding childbirth<br>HR (95%CI) |
| K52 | Other and unspecified noninfective gastroenteritis and colitis | 2.46 [0.74, 8.22] |
| M62 | Other disorders of muscle | 0.90 [0.49, 1.65] |
| N30 | Cystitis | 0.73 [0.42, 1.26] |
| O24 | Diabetes mellitus in pregnancy, childbirth, and the puerperium | 1.03 [0.61, 1.74] |
| O30 | Multiple gestation | 0.98 [0.60, 1.62] |
| O36 | Maternal care for other known or suspected fetal problems | 1.09 [0.82, 1.45] |
| O47 | False labor | 1.12 [0.84, 1.50] |
| O70 | Perineal laceration during delivery | 0.90 [0.70, 1.14] |
| O80 | Encounter for full-term uncomplicated delivery | 0.96 [0.79, 1.16] |
| O84 | Multiple births | 0.76 [0.38, 1.52] |
| O99 | Other maternal diseases classifiable elsewhere but complicating pregnancy, childbirth and the puerperium | 1.02 [0.77, 1.36] |
| R10 | Abdominal and pelvic pain | 1.06 [0.79, 1.42] |
| S02 | Fracture of skull and facial bones | 3.22 [0.85, 12.26] |
| S40 | Superficial injury of shoulder and upper arm | 1.69 [0.67, 4.30] |
| S60 | Superficial injury of wrist, hand and fingers | 1.19 [0.74, 1.94] |
| T14 | Injury of unspecified body region | 1.65 [0.79, 3.44] |
| Z00 | Encounter for general examination without complaint, suspected or reported diagnosis | 0.96 [0.46, 2.01] |
| Z29 | Encounter for other prophylactic measures | 0.74 [0.45, 1.20] |
| Z32 | Encounter for pregnancy test and childbirth and childcare instruction | 1.17 [1.01, 1.37]<sup>†</sup> |
| Z36 | Encounter for antenatal screening of mother | 0.94 [0.81, 1.08] |
| Z39 | Encounter for maternal postpartum care and examination | 1.00 [0.83, 1.22] |
| Z71 | Persons encountering health services for other counseling and medical advice, not elsewhere classified | 1.40 [0.89, 2.19] |

<sup>a</sup> model adjusted for maternal age at childbirth, total number of diagnoses in 24 months preceding childbirth, year of childbirth, and all the ICD-10 diagnostic categories presented in this table and chronic conditions: ICD-10: F10, F32, F33, F41, F43, F60, F99, G40, K07, K58
<sup>†</sup>: P-value <0.05; <sup>††</sup>: P-value <0.01; <sup>†††</sup>: P-value <0.001

